# The impact of education inequality on rheumatoid arthritis risk is mediated by smoking and body mass index: mendelian randomization study

**DOI:** 10.1101/2021.04.20.21254536

**Authors:** Sizheng Steven Zhao, Michael V Holmes, Jie Zheng, Eleanor Sanderson, Alice R Carter

## Abstract

**Objective:** To estimate the causal relationship between educational attainment – as a proxy for socioeconomic inequality – and risk of RA and quantify the roles of cigarette smoking and body mass index (BMI) as potential mediators.

**Methods:** Using the largest genome-wide association studies (GWAS), we performed a two-sample Mendelian randomization (MR) study of genetically predicted educational attainment (instrumented using 1265 variants from 766,345 individuals) and RA (14,361 cases, 43,923 controls). We used two-step MR to quantify the proportion of education’s effect on RA mediated by smoking exposure (as a composite index capturing duration, heaviness and cessation, using 124 variants from 462,690 individuals) and BMI (517 variants, 681,275 individuals), and multivariable MR to estimate proportion mediated by both factors combined.

**Results:** Each standard deviation (SD) increase in educational attainment (4.2 years of schooling) was protective of RA (OR 0.37; 95%CI 0.31, 0.44). Higher educational attainment was also protective for smoking exposure (β= -0.25 SD; 95%CI -0.26, -0.23) and BMI (β= -0.27 SD (^∼^1.3kg/m^2^); 95%CI -0.31, - 0.24). Smoking mediated 24% (95%CI 13%, 35%) and BMI 17% (95%CI 11%, 23%) of the total effect of education on RA. Combined, the two risk factors explained 47% (95%CI 11%, 82%) of the total effect.

**Conclusion:** Higher educational attainment has a protective effect on RA risk. Interventions to reduce smoking and excess adiposity at a population level may reduce this risk, but a large proportion of education’s effect on RA remains unexplained. Further research into other risk factors that act as potentially modifiable mediators are required.

**Key messages:**

1. Genetically predicted higher educational attainment – as a proxy for socioeconomic position – is protective for RA.
2. 24% of this effect was mediated by smoking behaviour and 17% by body mass index.
3. Efforts to reduce smoking and excess adiposity would help mitigate against socioeconomic inequalities in RA

## Introduction

Socioeconomic deprivation is recognised to be associated with increased risk of RA [1], but observational associations may have limited causal interpretation. Indices of local deprivation correlate poorly with individual socioeconomic position and are prone to ecological fallacy, while individual-level proxies such as income are often subject to reporting bias [2]. Furthermore, the causal direction is often difficult to establish; occupation, income and even area of residence (used to derive local deprivation indices) can each be influenced by work disability that can follow RA [3]. By contrast, educational attainment is largely determined in early life (predating, thus less likely influenced by, RA) and less likely to change over time (unlike income or occupation). Education is strongly correlated with employment, income and other later life measures of socioeconomic position, thereby serves as a good proxy [4].

The mechanism through which higher educational attainment protects against RA is not known; there have been no studies of causal intermediates, or mediators, to our knowledge. Education is intimately associated with smoking [5] and adiposity (e.g., measured using body mass index (BMI)) [6], but whether and to what extent these established risk factors explain the total effect of education on RA has not been investigated. Understanding the population-level implications of changes to smoking behaviour and BMI are important for reducing the effect of educational inequality on RA risk. Quantifying the proportion of the total effect unaccounted for may additionally highlight the need to study as yet undescribed intermediate factors. Such mediators may be more amenable to intervention, whereas efforts to improve educational opportunities across the population require intervention in early life and are beyond the scope of most clinical practices.

Mendelian randomisation (MR) mediation analysis can be used to address these unmet research needs. MR uses genetically predicted exposure levels to estimate its causal effect on the outcome. When instrumental variable assumptions are met, MR estimates are endowed with a causal interpretation and should not be subject to confounding, reverse causation, or measurement error. These strengths also apply to mediation analysis; for example, mediation analysis requires no unmeasured confounding between any of the exposure, mediator and outcome, which is difficult to achieve in traditional observational designs [7]. MR can also test reverse causation, e.g., whether RA or smoking influence educational attainment. Using two-sample MR, we aimed to investigate the effect of educational attainment on the risk of RA and quantify the roles of smoking and BMI as mediators.

## Methods

### GWAS Summary data

We obtained summary SNP-phenotype association data from genome-wide association studies (GWAS) of each respective phenotype (summarised in Table 1). Self-reported educational attainment was derived from the Social Science Genetic Association Consortium GWAS meta-analysis of years of schooling in 766,345 participants of European ancestry [8]. One standard deviation was equivalent to 4.2 years of additional schooling. BMI data were obtained from the Genetic Investigation of Anthropometric Traits consortium GWAS meta-analysis of 681,275 participants of European decent [9]. One standard deviation represents 4.8 kg/m^2^. Smoking was studied as “lifetime smoking exposure” in a GWAS of 462,690 participants of European ancestry in the UK Biobank. Rather than using binary smoking status, which may present methodological challenges [10], the smoking index takes into account smoking status (current/former/never), and exposure (duration/heaviness/cessation) among ever smokers, i.e., (1-0.5^smoking duration/half-life^)(0.5^time since cessation/half-life^)ln(cigarettes per day+1). This GWAS has been described in detail [11]. The index ranged from 0.007 (1 cigarette per day for 1 year) to 4.169 (currently smoking 140 a day and starting from age 11); one standard deviation increase in the smoking index is equivalent to, for example, an individual smoking 20 cigarettes a day for 15 years and stopping 17 years ago or an individual smoking 60 cigarettes a day for 13 years and stopping 22 years ago [11]. RA genetic associations were obtained from a GWAS meta-analysis of 14,361 RA cases (>90% met the 1987 American College of Rheumatology classification criteria) and 43,923 controls [12].

**Table 1:** Summary of each GWAS.

|  | Study population | Sample size | One standard deviation | No. of SNPs | r<sup>2</sup> | F |
| --- | --- | --- | --- | --- | --- | --- |
| Educational attainment [8] | Lee et al GWAS meta-analysis | 766,345 | 4.2 years | 1265 | 5.9% | 44.3 |
| BMI [9] | Yengo et al GWAS meta-analysis | 681,275 | 4.8 kg/m<sup>2</sup> | 517 | 5.3% | 73.7 |
| Lifetime smoking exposure*[11] | UK biobank | 462,690 | For example, smoking 20 cigarettes a day for 15 years and stopping 17 years ago | 124 | 0.5% | 41.4 |
| RA** [12] | Okada et al GWAS meta-analysis | 58,284 | n/a | 46 | n/a | n/a |
| Educational attainment [22] | Okbay et al GWAS | 293,723 | 3.7 years | 73 | 0.9% | 38.3 |
| Alcoholic drinks per week [26] | Liu et al GWAS | 335,394 | 9 additional drinks per week | 35 | 0.6% | 96.5 |
| Number of days/week of vigorous physical activity [25] | UK biobank | 440,512 | 1.95 days | 11 | 0.4% | 150 |
| Dietary protein | Meddens et al GWAS [27] | 268,922 | 102kcal<sup>#</sup> | 7 | 0.2% | 58.2 |
| Dietary fat |  | 268,922 | 270kcal<sup>#</sup> | 5 | 0.2% | 95.6 |
| Dietary carbohydrate |  | 268,922 | 334kcal<sup>#</sup> | 12 | 0.2% | 41.2 |
| *derived from smoking status (current, former, never), age at initiation in years, age at cessation in years and number of cigarettes smoked per day. **use in reverse MR of RA's effect on education. #One standard deviation increase in the context of mean total energy intake of 2064kcal. Dietary components as percentage of total energy intake measured using the food frequency questionnaire. n/a: effect allele frequency not available for RA to allow estimation of r2 and F statistic. GWAS, genome-wide association study; SNP, single nucleotide polymorphism; RA, rheumatoid arthritis; |  |  |  |  |  |  |

### Instrumental variable selection and data harmonisation

Genetic instruments for educational attainment were selected as the 1,265 independent genome-wide significant (p<5×10^−8^) SNPs that were not shared with variants instrumenting BMI or smoking. Instruments for BMI and smoking exposure were identified as the lead SNPs reaching genome-wide significance after removing SNPs in linkage disequilibrium (r^2^<0.001 or distance >10,000 kb) or shared with instruments for education. For all analyses, alleles were aligned to correspond to an increase in educational attainment. All effect alleles were checked to be on the forward strand. Where SNPs were absent in one of the exposure-outcome sets, SNPs in linkage disequilibrium (r^2^>0.9) were used as proxies. We calculated F statistics for each exposure in univariable MR, and conditional F statistics in MVMR, with F statistics >10 considered suggestive of adequate instrument strength [13].

### Effects of education on RA, smoking exposure and BMI

A graphical summary of analyses is given in Figure 1. First, we performed univariable two-sample MR to estimate the effect of educational attainment on RA (c in panel A) – referred to as the total effect – and effect of education on each mediator (a in panel B).

**Figure 1.**
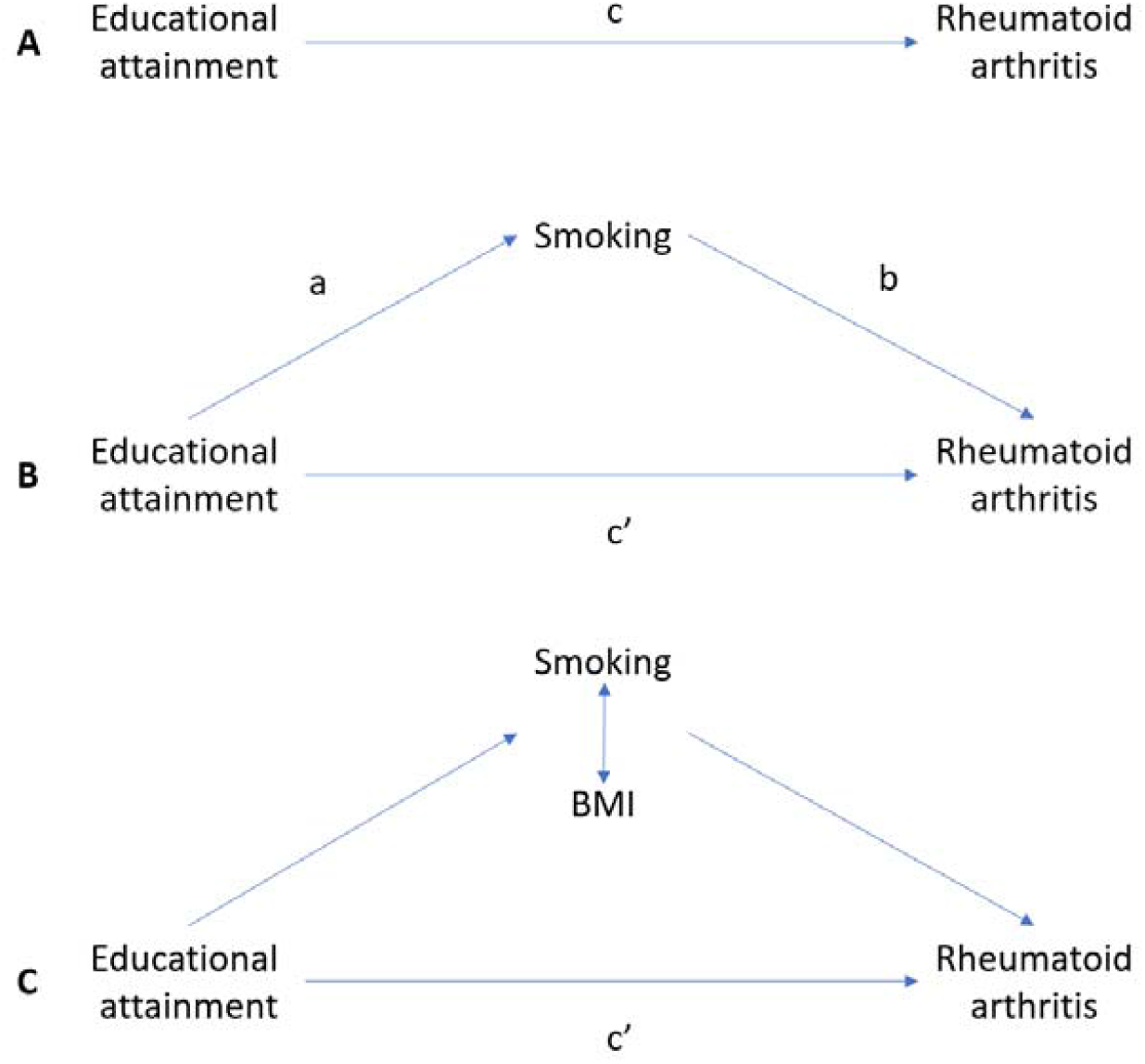
Summary of study design. **A**. The total effect of educational attainment (EA) on rheumatoid arthritis (RA), c, was derived using univariable MR (i.e., genetically predicted EA as exposure and RA as outcome). **B**. The total effect was decomposed into: 1) indirect effect using a two-step approach (where a is the total effect of EA on smoking, and b is the effect of smoking on RA adjusting for EA) and the product method (a*b) and 2) direct effect (c’ = c – a*b). The same process applied to mediation analysis of BMI. **C**. For mediation by both smoking and BMI combined, the indirect effect was derived using the difference method (c-c’). Proportion mediated was the indirect effect divided by the total effect.

We then used multivariable MR (MVMR) to estimate the effect of each mediator on RA (b in panel B), adjusting for education. For example, the effect of smoking on RA (b_1_) is: RA = b_1_*smoking + b_2_*education. MVMR was also used to model both mediators combined (panel C), i.e., RA = a*education + b_1_*smoking + b_2_*education.

### Decomposing mediated effects

The total effect of an exposure on an outcome can be decomposed into indirect (i.e., effect mediated through a causal intermediate) and direct effects (i.e., not through the mediator) [7]. The total effect of educational attainment on RA risk was decomposed into i) the direct effect of education on RA after adjusting for each mediator (c’ in panel C), and ii) the indirect effect of education through each mediator individually. The indirect effect of each mediator was derived using the product method; for example, the indirect effect of education on RA, through smoking, was obtained by multiplying the effect of education on smoking and the effect of smoking on RA (a*b in panel B).

To derive the indirect effect by smoking and BMI combined, the difference method was used (c-c’), where the direct effect, c’, was the effect of education adjusting for both smoking and BMI in an MVMR model.

For all mediators individually and combined, we quantified the proportion mediated by dividing indirect effect by the total effect. Confidence intervals were estimated using the delta method.

### Univariable and multivariable MR methods

We used the inverse-variance weighted method for the main univariable analysis, which combines results from each SNP using multiplicative random-effect meta-analysis [14]. Heterogeneity – a potential indicator of horizontal pleiotropy (and violation of MR assumptions) – was assessed using Cochran’s Q statistic. To test for potential bias from horizontal pleiotropy, we performed a series of sensitivity analyses using the weighted median [15] and mode-based estimators [16], MR-Egger regression [17] and MR-PRESSO (Pleiotropy RESidual Sum and Outlier) [18]. Each method relaxes certain MR assumptions such that a consistent effect across the multiple methods should be more robust against bias from horizonal pleiotropy (summarised in Table 2).

**Table 2:** summary of each MR method.

| MR method | Strengths and weaknesses |
| --- | --- |
| Inverse-variance weighted | A weighted mean of individual variant effects on the outcome, which provides an estimate equivalent to MR using individual-level data, assuming the genetic variants are uncorrelated. IVW has optimal statistical power, but assumes all variants are valid instruments. Estimates are biased if there is directional pleiotropy (when the average value of the pleiotropy distribution is non-zero) [14]. |
| MR-Egger | Quantifies directional pleiotropy and account for it to provide an unbiased estimate even if all the SNPs have pleiotropic effects. Requires the size of pleiotropic effects to be independent of the size of the variants' effects on the exposure (the InSIDE assumption), which is not verifiable. It is sensitive to outliers and less efficient (result in wide confidence intervals) [17]. |
| Weighted median | Robust to outliers; unbiased estimate when up to half of the SNPs violate the instrumental variable assumptions. May be less efficient [15]. |
| Mode-based estimation | Robust to outliers; assumes SNPs in the largest cluster are valid instruments and provide an unbiased estimate even if most other SNPs are invalid [16]. |
| MR-PRESSO | Identifies and removes potentially pleiotropic outliers. High false positive rate when there are several invalid IVs [18]. |
| Multivariable MR | extension of univariable MR that estimates the effect of two or more exposures on an outcome, where a secondary exposure acts can act as a confounder, a mediator, a pleiotropic pathway or a collider [19]. It relies on knowledge of the covariance between the effect of the SNP on each exposure, which are not always available in conventional GWAS results. |
| IV, instrumental variable; MR-PRESSO, Mendelian Randomization Pleiotropy RESidual Sum and Outlier. |  |

For MVMR, we used the inverse-variance weighted method, with MR-Egger as sensitivity analysis [19]. The pairwise covariance between SNP associations was assumed to be zero in the primary analysis. We tested this assumption using a range of covariance values. Conditional instrument strength was quantified using the modified F-statistic and heterogeneity was assessed using modified Cochran’s Q statistic [19].

### Sensitivity analyses

First, we tested for potential mis-specification of the exposure (i.e., whether SNPs influence the exposure first and then the outcome) using Steiger filtering [20]. We also tested potential for reverse causation, that is, using genetic instruments for RA, smoking exposure and BMI to examine their effects on educational attainment; genetic instruments were chosen using the same approach as above. Second, we tested potential bias from overlapping samples [21] (GWAS meta-analyses for education, smoking and BMI all contain UK Biobank participants) using an earlier GWAS of educational attainment without UK Biobank participants [22]. Third, to address weak conditional instrument strength in MVMR analyses including smoking we: i) restricted analyses to the most strongly associated SNPs for each exposure where conditional instrument strength was weak (this reduces bias from weak instruments but also reduces precision) and ii) using the weak instrument robust estimator [23].

Although smoking and BMI are the leading modifiable risk factors for RA, prior studies have also proposed several related lifestyle factors as risk factors [24]. We additionally considered physical activity (number of days per week of vigorous physical activity lasting >10 minutes) [25], alcohol consumption (drinks per week) [26], and dietary composition (self-reported relative fat, protein, carbohydrate intake) [27] as risk factors for RA. All analyses were performed in R using the TwoSampleMR and MVMR packages [19,28].

## Results

Genetic instruments for educational attainment explained 5.9% of its variance, with an univariable F statistic of 38. The variance explained by, and F statistic for, SNPs instrumenting smoking exposure were 0.5% and 37, and BMI 5.3% and 74.

### Effects of education on RA, BMI and smoking behaviour

For each standard deviation (4.2 years) increase in educational attainment, the relative odds of RA were 63% lower (OR 0.37; 95%CI 0.31, 0.44). Higher educational attainment was associated with lower smoking exposure (β= -0.25; 95%CI -0.26, -0.23) and lower BMI (β= -0.27; 95%CI -0.31, -0.24; i.e., one SD increase in education was associated with 1.3kg/m^2^ lower BMI).

### Effect of BMI and smoking behaviour on RA

In univariable MR, each standard deviation increase in smoking exposure (OR 2.13; 1.25, 3.62) or in BMI (OR 1.14; 0.95, 1.36) led to a higher relative odds of RA (Figure 2).

**Figure 2.**
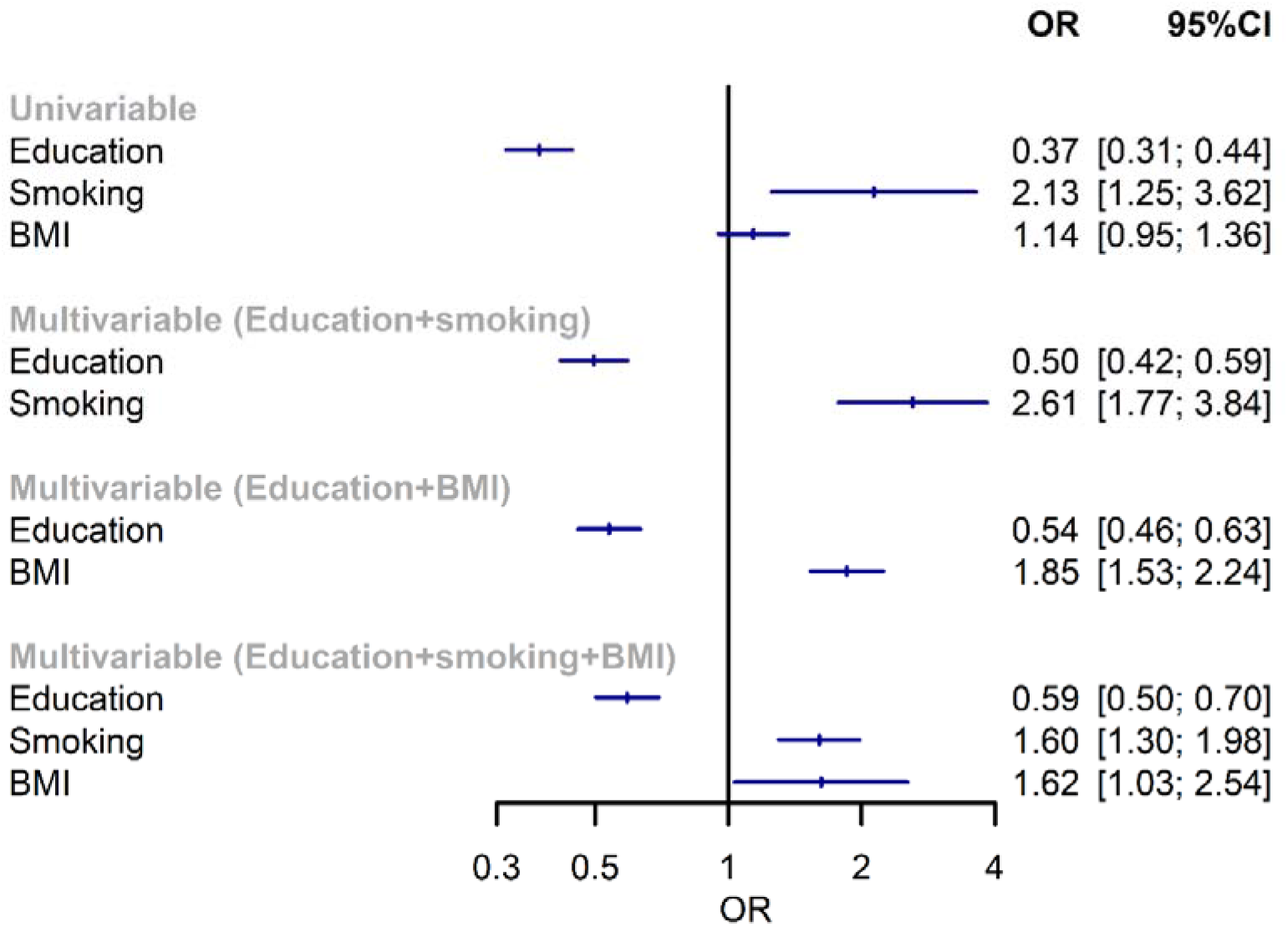
Effect of one standard deviation increase in each exposure on odds of rheumatoid arthritis in uni- and multivariable models. BMI, body mass index; OR, odds ratio; CI, confidence interval.

There were bidirectional positive effects between smoking and BMI: smoking exposure increased BMI (β=0.59; 0.34, 0.84), while BMI increased smoking exposure (β=0.11; 0.10, 0.13)

### Mediation by smoking and BMI behaviour

In the MVMR analysis of education-smoking-RA, the conditional F statistics for educational attainment and smoking exposure were 25 and 8.4, respectively. The direct effect of educational attainment on RA was OR 0.50 (95%CI 0.42, 0.59) after accounting for smoking (Figure 2). The direct effect of smoking on RA was OR 2.61 (95%CI 1.77, 3.84) after accounting for education. The proportion mediated by smoking was 24% (95%CI 13%, 35%) (Figure 3).

**Figure 3.**
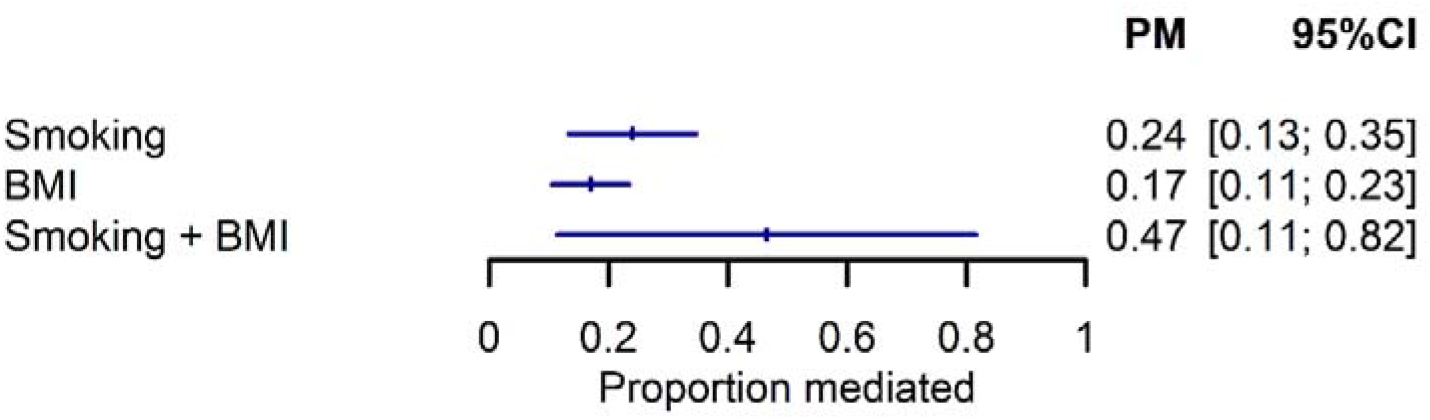
Estimate of the effect of education on rheumatoid arthritis explained by each mediator and by both combined. BMI, body mass index; OR, odds ratio; CI, confidence interval.

Conditional F statistics for educational attainment and BMI were 29 and 20, respectively, in the MVMR analysis of education-BMI-RA. The direct effect of educational attainment on RA was OR 0.54 (95%CI 0.46, 0.63) after accounting for BMI (Figure 2). After accounting for education, the direct effect of BMI on RA was OR 1.85 (1.53, 2.24). The proportion mediated by BMI was 17% (95%CI 11%, 23%) (Figure 3).

When both smoking and BMI were entered into the MVMR model, conditional instrument strength was further reduced (education 18, smoking 5.8, BMI 10). Effect sizes for education (OR 0.59; 95%CI 0.50, 0.70), BMI (OR 1.60 95%CI 1.30, 1.98) and smoking exposure (OR 1.62; 95%CI 1.03, 2.54) were attenuated (Figure 2). Combined, BMI and smoking mediated 47% (95%CI 11%, 82%) of the effect of education on RA (Figure 3).

### Sensitivity analyses

MR sensitivity methods had reduced precision, but generally did not change the causal direction of estimates (Supplementary Tables S1 and S2, Figures S1 to S5). There was significant heterogeneity for all MR analyses (Supplementary Table S3), but no evidence of directional pleiotropy (Supplementary Table S4). There was no evidence of reverse causation according to Steiger directionality test (Supplementary Table S5) or reverse MR (Supplementary Table S6).

Using a smaller education GWAS without UK Biobank, the total effect of education (where one SD was 3.6 years of schooling) on RA was similar to the primary analysis (Supplementary Figure S6). The proportion mediated by smoking (28%; 95%CI 10%, 46%) and BMI (29%; 95%CI 8%, 49%) were similar but lacked precision; estimate for BMI and smoking combined was 45% with CI out of bounds (Supplementary Figure S7). Restricting to the 10% most strongly associated SNPs for each exposure in the MVMR models of education-smoking (conditional F = 48 and 14, respectively) and education-smoking-BMI (F = 17, 12, 20) lead to point estimates within confidence intervals of the primary analysis, but with reduced precision (Supplementary Figure S6). Effect estimates were again similar using the weak instrument robust estimator irrespective of covariance (Supplementary Figure S6). There was no evidence of a total effect of alcohol consumption, physical activity, or dietary composition on RA risk in univariable MR (Supplementary Table S7), thus mediation was not tested. All SNPs and proxies used are shown in Supplementary Tables S8.

## Discussion

Genetically predicted higher educational attainment led to lower relative odds of RA (63% lower for every additional 4.2 years of education). One quarter of this effect was mediated through smoking, and 17% through BMI. In this study, these two leading risk factors for RA accounted for 47% of the total effect of education, suggesting that over half of the effect of education on RA remains unexplained.

This is the first application of MR mediation analysis to study mediators of education and RA risk. The protective effect of higher educational attainment is congruent with findings from traditional observational designs. Each of the following studies also used education as a proxy of wider socioeconomic inequality. Bergtsson et al showed that Swedish patients with university (vs no university) degree had 29% lower relative risk (inverse of the reported RR 1.4) of RA [29]. The effect size was largely unchanged (RR 1.5) when additionally adjusting for smoking status. This small change from unadjusted (total) to adjusted (direct) effects may be explained by imprecision with which smoking exposure was measured. Pedersen et al showed those with “long term advanced studies” (formal education >4 years) had lower odds of RA compared to those with no formal education (OR 0.30; 0.18, 0.51); the effect size was reduced (OR 0.43; 0.24, 0.76) after adjusting for smoking, BMI, physical activity [30]. These results are difficult to interpret since years of schooling was also included in models. Both studies were case-control in design with low response rates that differed between the groups – a key source of bias. Neither formally considered a mediation approach to quantify the relative contribution of different risk factors.

Higher educational attainment has been shown to be protective for a range of health outcomes, the effect sizes of which attenuates with adjustment for (i.e., suggestive of mediation by) smoking and BMI [31]. For example, smoking mediated 28% and BMI mediated 18% of education’s effect on myocardial infarction [32]. This suggests that public health interventions to reduce smoking and excess adiposity would have widespread benefits on many prevalent comorbidities in RA that drive mortality and additional morbidity.

Our results also suggest that education (and potentially the socioeconomic deprivation that it proxies) may be associated with other important environmental risk factors that increase RA risk. In our analysis, over half of the effect of education remains unaccounted for. Alcohol consumption, physical activity and dietary composition did not appear to have meaningful causal effects on RA risk in our sensitivity analyses. These null results contrast with a host of observational studies that show associations with RA risk, which may reflect unmeasured confounding, measurement error or other biases [33–35]. For example, the allegedly protective properties of alcohol could be due to reverse causation [36]. Null results may also be explained by the difficulty in accurately measuring these exposures in GWASs.

Although educational attainment is commonly used as a proxy for socioeconomic position, it is important to note that they are not interchangeable. Socioeconomic position encapsulates many more factors than education; equally, education may affect health outcomes through mechanisms independent of socioeconomic position. Other measures of socioeconomic position may produce different results to those found here. Genetic instruments for education often do not have a clear biological basis and may instead causally influence related traits closely correlated with education (e.g., other proxies for socioeconomic position).

A key strength of this study is the ability of MR to provide causal estimates. In the context of mediation, MR provides further robustness to non-differential measurement error in the mediator [7]. We used lifetime smoking exposure, which captures dimensions of smoking behaviour overlooked when studying smoking status [11]. There were also limitations. A main source of potential bias in MR studies is horizontal pleiotropy; we examined this using a myriad of MR methods that provided consistent results to the main analysis. We were not able to examine potential for exposure-mediator interaction, e.g., estimates would be biased if smoking behaviour interacts with causal effect of education on RA risk. The current analysis assumes a linear effect of education on each outcome; it is nevertheless a valid test of the causal null hypothesis even should this assumption not hold [37]. Prior observational studies of deprivation and education showed a particularly strong association with seropositive RA [30]; we were not able to examine serostatus in the current study. Inferences drawn from our results apply to RA onset, and may not be extrapolatable to RA prognosis (indeed, MR studies of disease progression may be subject to additional biases [38]). Instrumental variable methods (including MR) estimate the ‘local average treatment effects’ not population average effects; here it is the average effect of education for individuals whose educational attainment was increased by the SNPs (i.e., a subgroup). This further requires the direction of SNP-effects on education, and indeed the SNP-effects on each mediator, to be in the same direction for all individuals (‘monotonicity’). Neither the subgroup proportion nor monotonicity can be empirically verified. Lastly, MR may not capture time-varying nature of the exposures; a snapshot BMI measurement may not fully represent BMI over the life course [39]. This is particularly relevant to genetic instruments for physical activity, diet and alcohol intake, which we considered in sensitivity analyses.

In conclusion, we showed a protective effect of higher educational attainment on RA risk; interventions to reduce smoking and excess adiposity at a population level may reduce much of this risk for RA. This is particularly relevant for those with high risk of developing RA (e.g., those with strong family history). However, the majority of education’s effect on RA remain unexplained. Broader efforts to improve socioeconomic inequalities and access to education are required, as well as further research into other environmental risk factors that act as potentially modifiable mediators of socioeconomic deprivation.

## Data Availability

Summary statistics are available from each consortia (details in Table 1) or via the MR-Base platform (https://gwas.mrcieu.ac.uk/).

## Contribution

SSZ analysed the data and wrote the manuscript with significant input from all co-authors.

## Ethics declarations

Participating studies of respective GWAS meta-analyses have received prior approval by relevant institutional review boards, and informed consent was obtained from each study participant (details in Table 1). The current study used publicly available summary statistics data; therefore, no additional ethics approval was required.

## References

1. Calixto O-J, Anaya J-M. Socioeconomic status. The relationship with health and autoimmune diseases. Autoimmunity Reviews 2014;13:641–54.

2. Verstappen SMM. The impact of socio-economic status in rheumatoid arthritis. Rheumatology (Oxford) 2017;56:1051–2.

3. Doeglas D, Suurmeijer T, Krol B, Sanderman R, van Leeuwen M, van Rijswijk M. Work disability in early rheumatoid arthritis. Ann Rheum Dis 1995;54:455–60.

4. Galobardes B, Shaw M, Lawlor DA, Lynch JW, Davey Smith G. Indicators of socioeconomic position (part 1). J Epidemiol Community Health 2006;60:7–12.

5. Qian Y, Zhang L, Wu DJH, Xie Z, Wen C, Mao Y. Genetic predisposition to smoking is associated with risk of rheumatoid arthritis: a Mendelian randomization study. Arthritis Research & Therapy 2020;22:44.

6. Tang B, Shi H, Alfredsson L, Klareskog L, Padyukov L, Jiang X. Obesity-related traits and the development of rheumatoid arthritis - evidence from genetic data. Arthritis Rheumatol 2021 Feb;73(2):203–211

7. Carter AR, Sanderson E, Hammerton G, Richmond RC, Smith GD, Heron J, et al. Mendelian randomisation for mediation analysis: current methods and challenges for implementation. bioRxiv 2020;835819.

8. Lee JJ, Wedow R, Okbay A, Kong E, Maghzian O, Zacher M, et al. Gene discovery and polygenic prediction from a 1.1-million-person GWAS of educational attainment. Nat Genet 2018;50:1112–21.

9. Yengo L, Sidorenko J, Kemper KE, Zheng Z, Wood AR, Weedon MN, et al. Meta-analysis of genome-wide association studies for height and body mass index in ∼700000 individuals of European ancestry. Hum Mol Genet 2018;27:3641–9.

10. Burgess S, Labrecque JA. Mendelian randomization with a binary exposure variable: interpretation and presentation of causal estimates. Eur J Epidemiol 2018;33:947–52.

11. Wootton RE, Richmond RC, Stuijfzand BG, Lawn RB, Sallis HM, Taylor GMJ, et al. Evidence for causal effects of lifetime smoking on risk for depression and schizophrenia: a Mendelian randomisation study. Psychol Med 2020;50:2435–43.

12. Okada Y, Wu D, Trynka G, Raj T, Terao C, Ikari K, et al. Genetics of rheumatoid arthritis contributes to biology and drug discovery. Nature 2014;506:376–81.

13. Burgess S, Thompson SG, CRP CHD Genetics Collaboration. Avoiding bias from weak instruments in Mendelian randomization studies. International Journal of Epidemiology 2011;40:755–64.

14. Burgess S, Butterworth A, Thompson SG. Mendelian randomization analysis with multiple genetic variants using summarized data. Genet Epidemiol 2013;37:658–65.

15. Bowden J, Davey Smith G, Haycock PC, Burgess S. Consistent Estimation in Mendelian Randomization with Some Invalid Instruments Using a Weighted Median Estimator. Genet Epidemiol 2016;40:304–14.

16. Hartwig FP, Davey Smith G, Bowden J. Robust inference in summary data Mendelian randomization via the zero modal pleiotropy assumption. International Journal of Epidemiology 2017;46:1985–98.

17. Bowden J, Davey Smith G, Burgess S. Mendelian randomization with invalid instruments: effect estimation and bias detection through Egger regression. International Journal of Epidemiology 2015;44:512–25.

18. Verbanck M, Chen C-Y, Neale B, Do R. Detection of widespread horizontal pleiotropy in causal relationships inferred from Mendelian randomization between complex traits and diseases. Nat Genet 2018;50:693–8.

19. Sanderson E, Davey Smith G, Windmeijer F, Bowden J. An examination of multivariable Mendelian randomization in the single-sample and two-sample summary data settings. International Journal of Epidemiology 2019;48:713–27.

20. Hemani G, Tilling K, Smith GD. Orienting the causal relationship between imprecisely measured traits using GWAS summary data. PLOS Genetics 2017;13:e1007081.

21. Burgess S, Davies NM, Thompson SG. Bias due to participant overlap in two-sample Mendelian randomization. Genetic Epidemiology 2016;40:597–608.

22. Okbay A, Beauchamp JP, Fontana MA, Lee JJ, Pers TH, Rietveld CA, et al. Genome-wide association study identifies 74 loci associated with educational attainment. Nature 2016;533:539–42.

23. Sanderson E, Spiller W, Bowden J. Testing and Correcting for Weak and Pleiotropic Instruments in Two-Sample Multivariable Mendelian Randomisation. bioRxiv 2020;2020.04.02.021980.

24. Smolen JS, Aletaha D, Barton A, Burmester GR, Emery P, Firestein GS, et al. Rheumatoid arthritis. Nature reviews. Disease primers 2018;4:18001.

25. Trait: Duration of vigorous activity - IEU OpenGWAS project [Internet]. [cited 2021 Mar 1]; Available from: https://gwas.mrcieu.ac.uk/datasets/ukb-b-13932/

26. Liu M, Jiang Y, Wedow R, Li Y, Brazel DM, Chen F, et al. Association studies of up to 1.2 million individuals yield new insights into the genetic etiology of tobacco and alcohol use. Nat Genet 2019;51:237–44.

27. Meddens SFW, Vlaming R de, Bowers P, Burik CAP, Linnér RK, Lee C, et al. Genomic analysis of diet composition finds novel loci and associations with health and lifestyle. Molecular Psychiatry 2020;1–14.

28. Walker VM, Davies NM, Hemani G, Zheng J, Haycock PC, Gaunt TR, et al. Using the MR-Base platform to investigate risk factors and drug targets for thousands of phenotypes. Wellcome Open Res. 2019; 4: 113.

29. Bengtsson C, Nordmark B, Klareskog L, Lundberg I, Alfredsson L, EIRA Study Group. Socioeconomic status and the risk of developing rheumatoid arthritis: results from the Swedish EIRA study. Ann Rheum Dis 2005;64:1588–94.

30. Pedersen M, Jacobsen S, Klarlund M, Frisch M. Socioeconomic status and risk of rheumatoid arthritis: a Danish case-control study. J Rheumatol 2006;33:1069–74.

31. Yuan S, Xiong Y, Michaëlsson M, Michaëlsson K, Larsson SC. Health-related effects of education level: a Mendelian randomization study. medRxiv 2020;2020.02.01.20020008.

32. Carter AR, Gill D, Davies NM, Taylor AE, Tillmann T, Vaucher J, et al. Understanding the consequences of education inequality on cardiovascular disease: mendelian randomisation study. BMJ 2019;365:1855.

33. Liu X, Tedeschi SK, Lu B, Zaccardelli A, Speyer CB, Costenbader KH, et al. Long-Term Physical Activity and Subsequent Risk for Rheumatoid Arthritis Among Women: A Prospective Cohort Study. Arthritis & Rheumatology 2019;71:1460–71.

34. Giuseppe DD, Alfredsson L, Bottai M, Askling J, Wolk A. Long term alcohol intake and risk of rheumatoid arthritis in women: a population based cohort study. BMJ 2012;345:e4230.

35. Cutolo M, Nikiphorou E. Don’t neglect nutrition in rheumatoid arthritis! RMD Open 2018;4:e000591.

36. Baker JF, England BR, Mikuls TR, Hsu JY, George MD, Pedro S, et al. Changes in Alcohol Use and Associations with Disease Activity, Health Status, and Mortality in Rheumatoid Arthritis. Arthritis Care & Research 2020 Mar;72(3):301–308

37. Burgess S, Davies NM, Thompson SG, EPIC-InterAct Consortium. Instrumental variable analysis with a nonlinear exposure-outcome relationship. Epidemiology 2014;25:877–85.

38. Paternoster L, Tilling K, Davey Smith G. Genetic epidemiology and Mendelian randomization for informing disease therapeutics: Conceptual and methodological challenges. PLoS Genet 2017;13:e1006944.

39. Labrecque JA, Swanson SA. Commentary: Mendelian randomization with multiple exposures: the importance of thinking about time. International Journal of Epidemiology 2020;49:1158–62.

