## Supplementary Figure for "The impact of education inequality on rheumatoid arthritis risk is mediated by smoking and body mass index: mendelian randomization study"

**Supplementary Figure S1. Scatter plot for the effect of education on rheumatoid arthritis.**

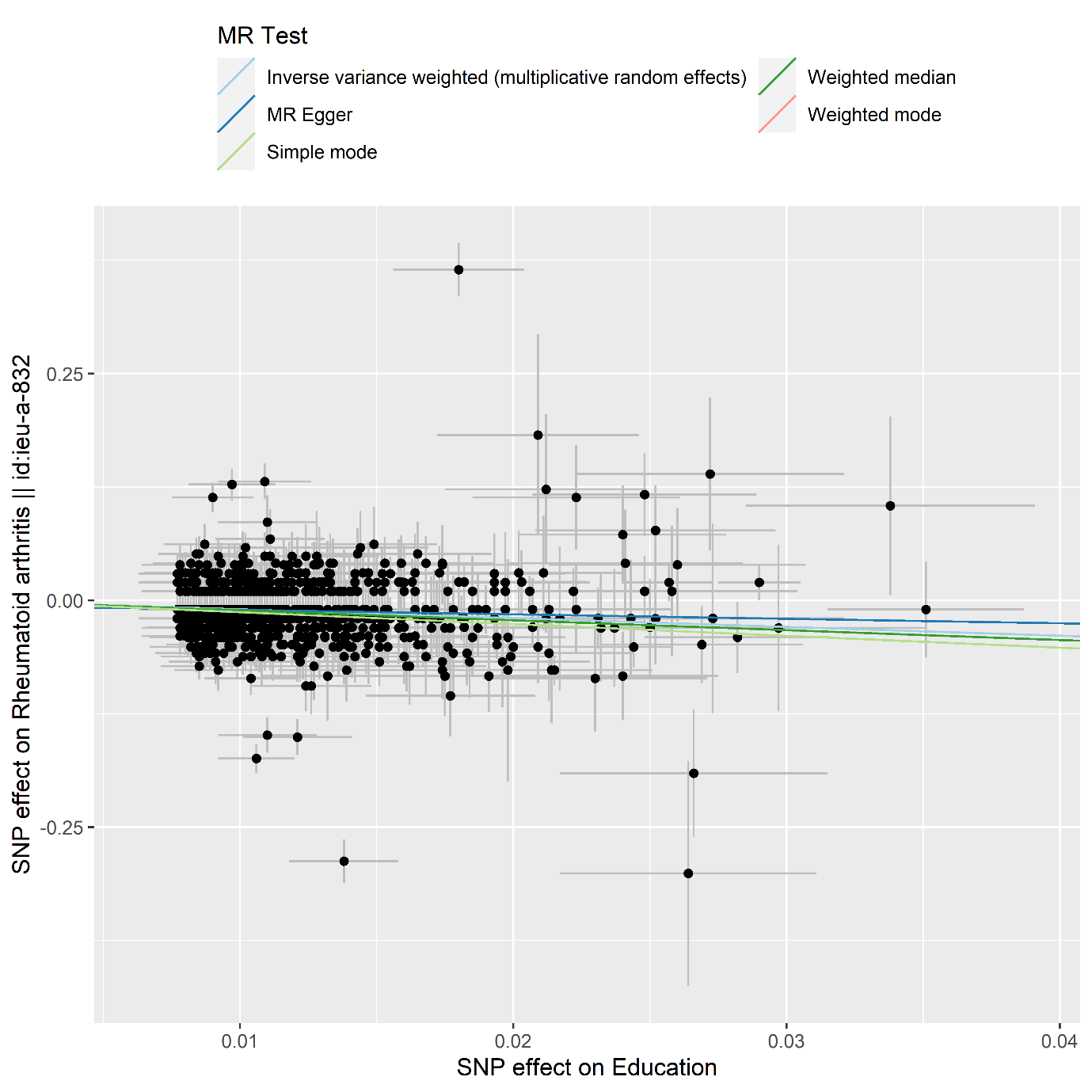

**Supplementary Figure S2. Scatter plot for the effect of smoking exposure on rheumatoid arthritis.**

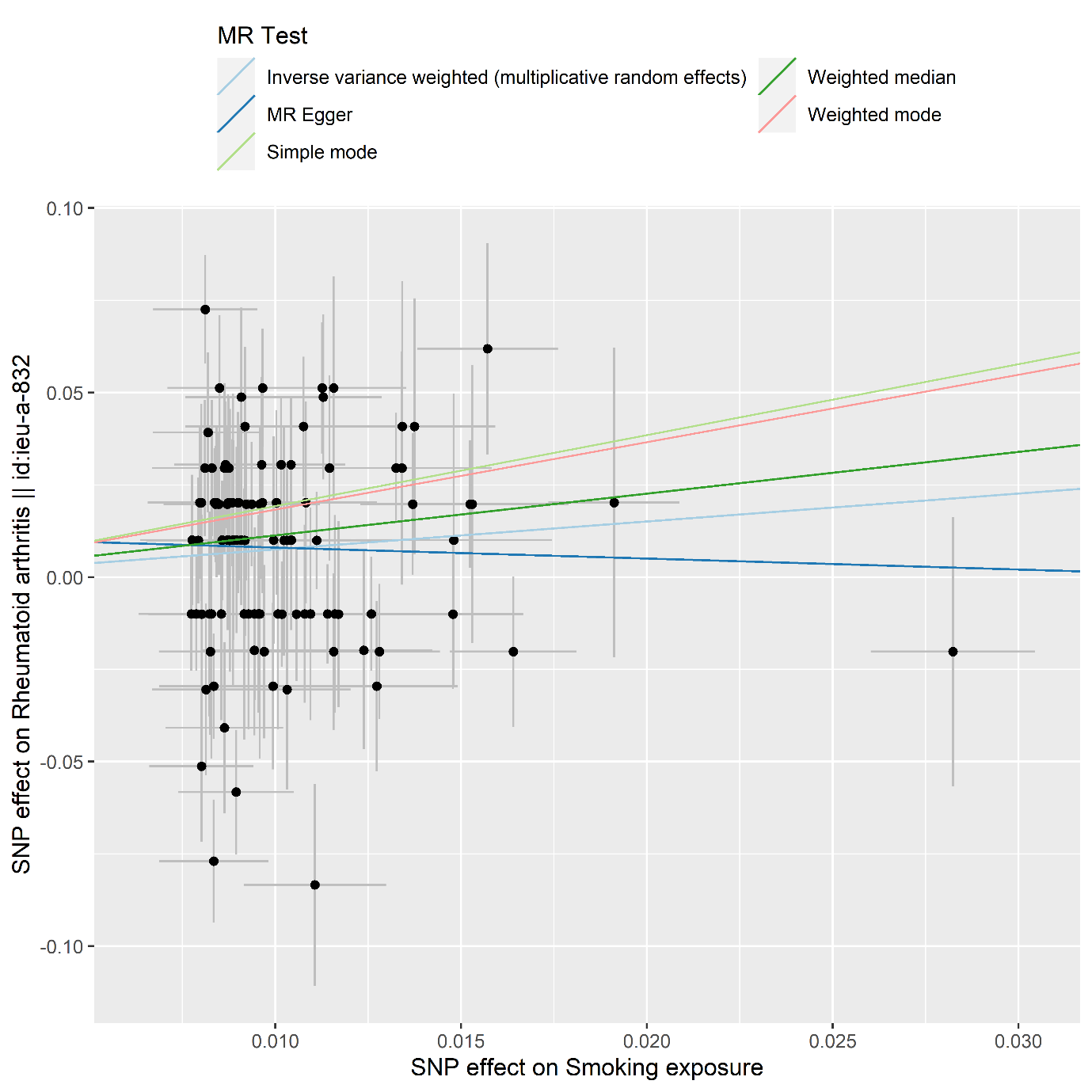

**Supplementary Figure S3. Scatter plot for the effect of BMI on rheumatoid arthritis.**

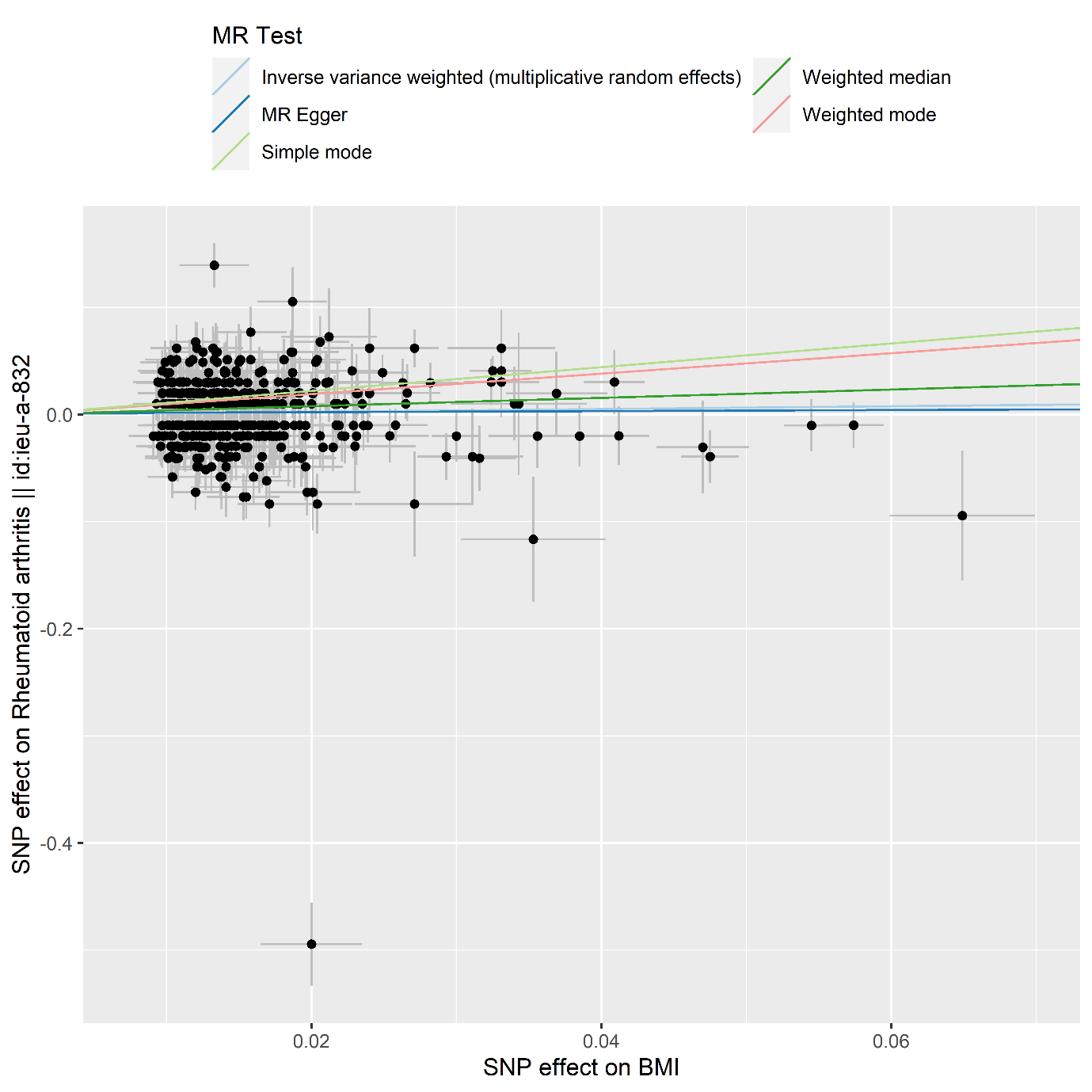

**Supplementary Figure S4. Scatter plot for the effect of education on smoking exposure.**

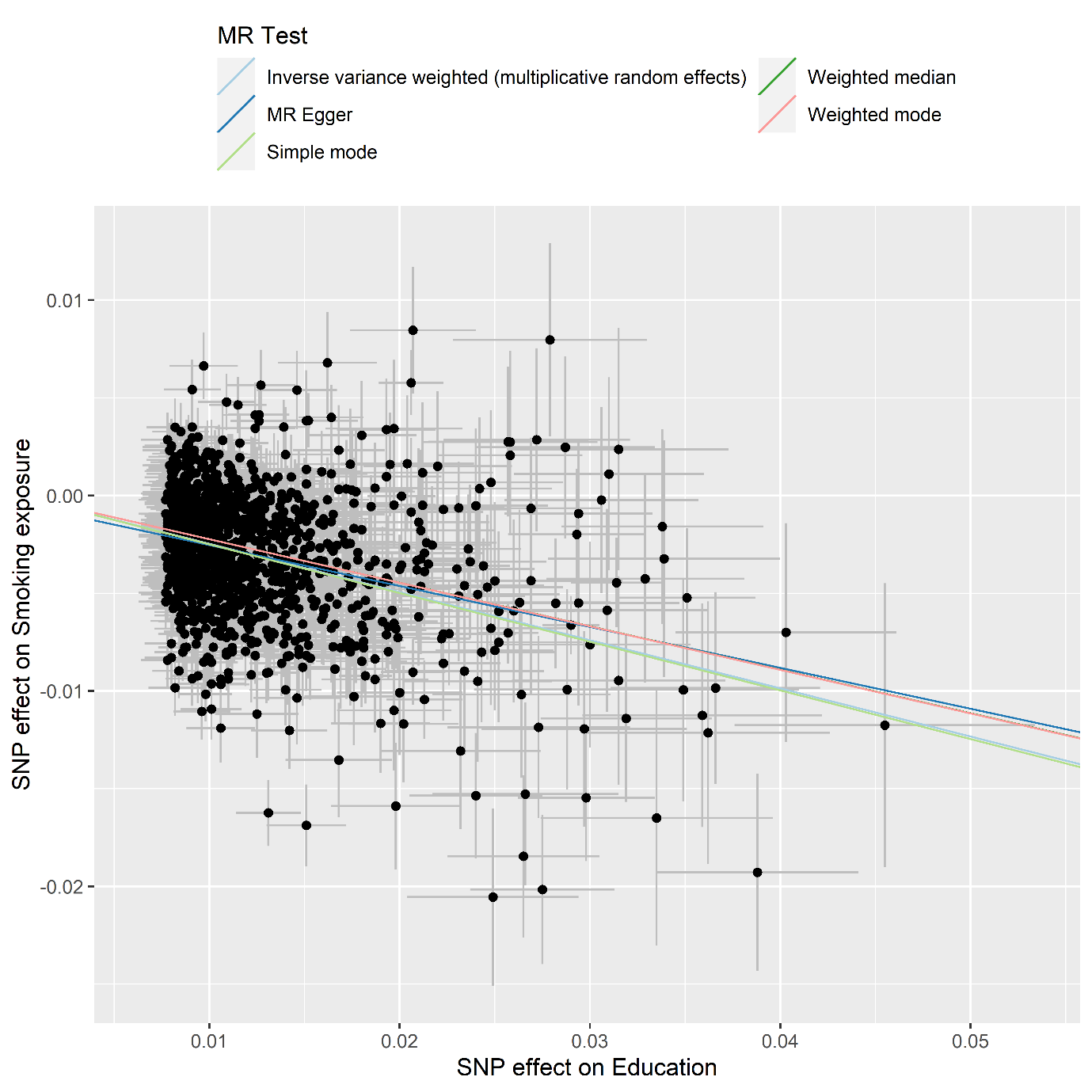

**Supplementary Figure S5. Scatter plot for the effect of education on smoking exposure.**

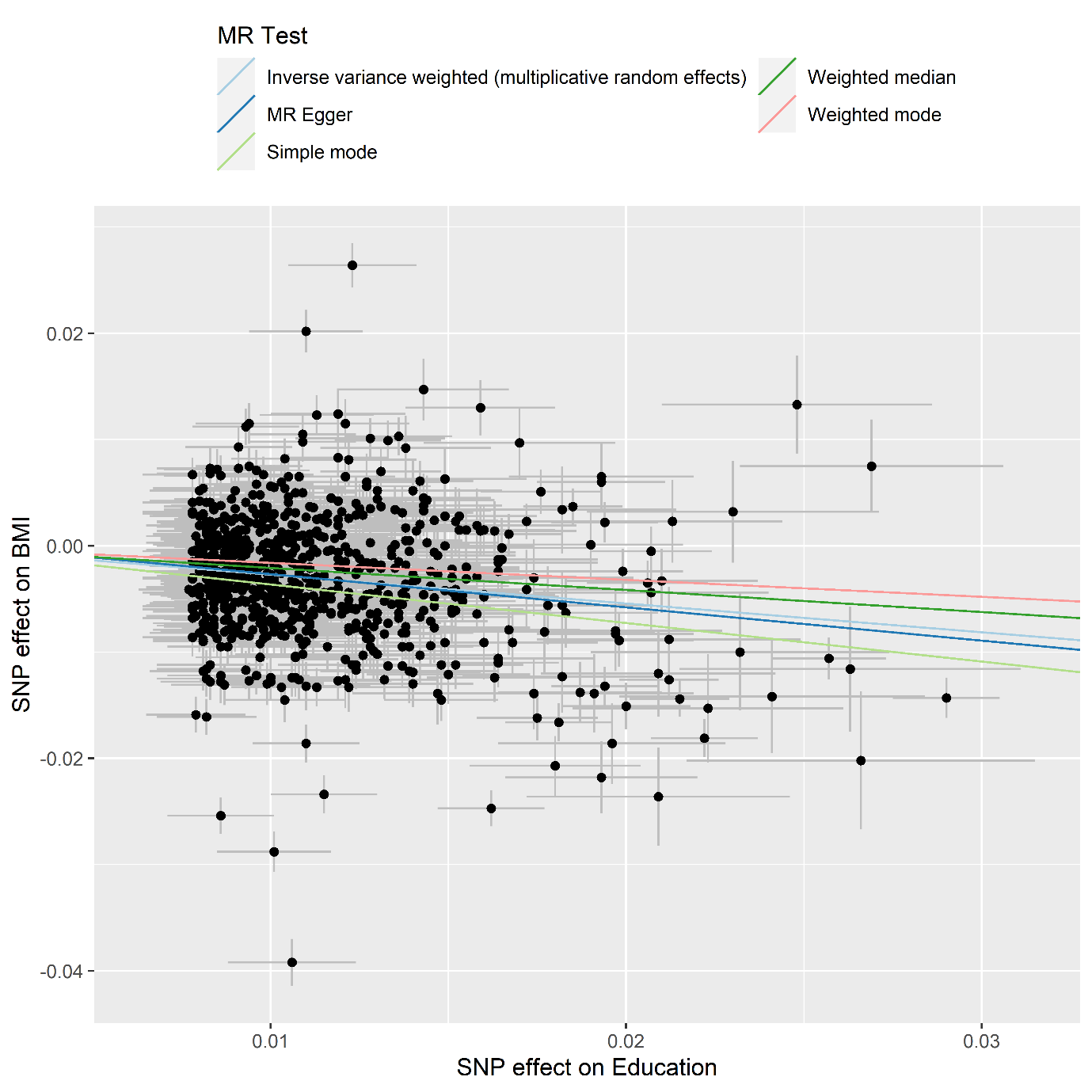

**Supplementary Figure S6. Forest plot summarising sensitivity analysis results.**

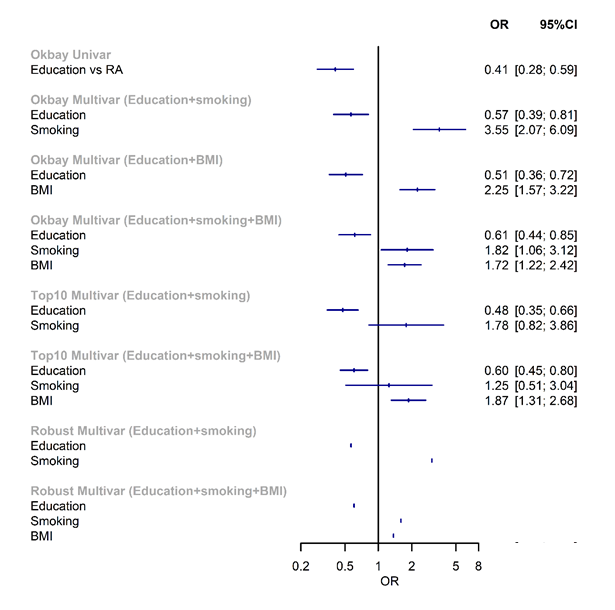

Okbay: primary analysis repeated using a small GWAS by Okbay et al that does not have sample overlap with UK biobank. Top 10: sensitivity analysis restricting to the top 10% most SNPs most strongly associated with each exposure, for MVMR with conditional F statistic <10. Robust: weak instrument robust method; note that confidence intervals were not estimated due to computational burden.

**Supplementary Figure S7. Forest plot summarising proportion mediated in the primary and sensitivity analysis using the smaller education GWAS by Okbay et al.**
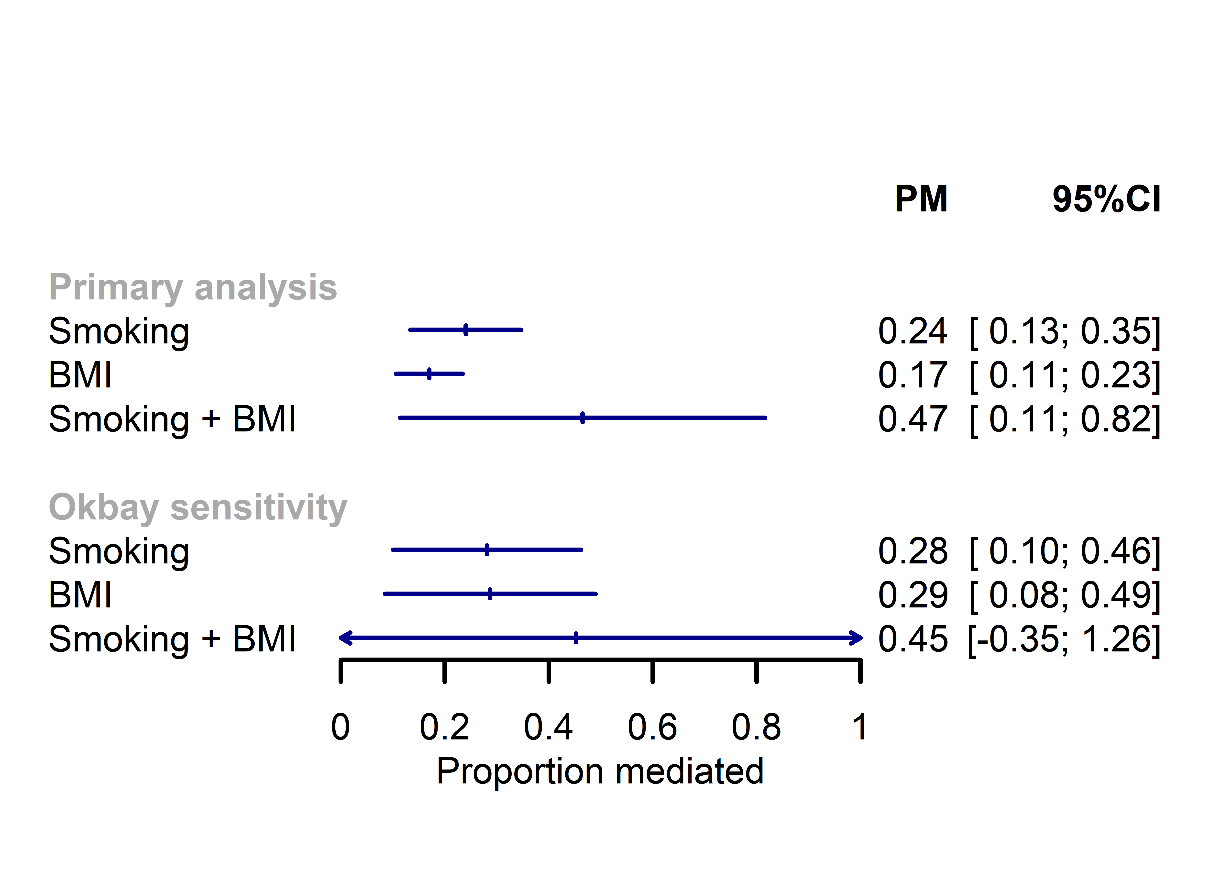
